# Whole-Lung CT Radiomics-Based Machine Learning Classification of Nontuberculous Mycobacterial Lung Disease Across Geographically Distinct Cohorts

**DOI:** 10.64898/2026.06.29.26356713

**Authors:** Anusha Kanagala, Bryan Garcia, Taru S. Dutt, Stéphanie M. Aguilera, Abhiraj S. Pudhota, Dhiral D. Panjwani, Nikhil Dukkipati, Amit Gaggar, Threnesan Naidoo, Leon Jololian, Surya P. Bhatt, Camilla Margaroli, Sandeep Bodduluri

## Abstract

**BACKGROUND:** Nontuberculous mycobacterial lung disease (NTM-LD) is highly heterogenous, geographically and etiologically, hindering effective timely identification. Prior CT radiomics studies require manual segmentation of pathology. We developed a whole-lung CT radiomics-based machine learning approach and identified common features across two geographically distinct NTM-LD cohorts.

**STUDY DESIGN AND METHODS:** 1,300 chest CT scans from China (871 TB; 429 NTM, Dataset 1) and 173 independent NTM cohort from UAB, US. Whole-lung regions were automatically segmented on each scan, and 85 quantitative radiomic features were extracted using a standardized image-processing pipeline. We evaluated two frameworks to assess model performance and generalizability: (1) training on Dataset 1 with external validation on Dataset 2, and (2) training on the combined cohort. Linear discriminant analysis (LDA) was used as the primary classification method. Cross-cohort concordance analysis was performed to evaluate the reproducibility of radiomic features across datasets.

**RESULTS:** In Scenario 1, the LDA classifier trained on Dataset 1 achieved an AUC of 0.79 (95% CI, 0.73–0.84) with high specificity (0.91). On the external UAB cohort, the model achieved an AUC of 0.94 (95% CI, 0.90–0.97). In Scenario 2, the combined cohort model achieved an AUC of 0.81 (95% CI, 0.76–0.85) with improved sensitivity (0.61) and precision (0.82). Feature importance analysis identified 16 features consistently ranked among the top 20 in both scenarios, predominantly texture-based descriptors reflecting distinct parenchymal patterns between myco-bacterial species.

**CONCLUSION:** Whole-lung CT radiomics enables interpretable NTM-LD classification across geographically distinct populations without manual annotation. Suggesting population-independent parenchymal signatures of NTM-LD.

## INTRODUCTION

Nontuberculous mycobacterial lung disease (NTM-LD) and pulmonary tuberculosis (PTB) represent distinct clinical entities requiring fundamentally different treatment and management approaches, yet their differentiation poses substantial diagnostic challenges.^1,2^ At present, distinguishing NTM-LD from PTB often relies on sputum-based microbiologic testing, including smear microscopy, mycobacterial culture, and nucleic acid amplification tests such as Xpert MTB/RIF. However, culture remains time-consuming because mycobacteria require weeks to grow, and molecular assays can detect DNA from nonviable bacilli, limiting their ability to distinguish active infection from residual bacterial material. The global incidence of NTM-LD has increased substantially over recent decades, with prevalence now exceeding that of TB in many developed nations including the United States, while TB remains a leading cause of infectious disease mortality worldwide.^3,4^ Both conditions manifest with overlapping clinical presentations and radiologic findings on chest CT imaging, including nodular infiltrates, cavitary lesions, bronchiectasis, and tree-in-bud patterns.^5,6^ Early differentiation is essential not only for appropriate antimicrobial selection but also for infection control measures, as faster identification can reduce community exposure to PTB and avoid unnecessary airborne isolation in patients with NTM-LD. Additional challenges in the timely clinical diagnosis of NTM-LD are posed by the microbiological diversity of NTM, which includes more than 150 species, and the etiological differences of underlying lung diseases acting as predisposing factors for NTM-LD (PMID: 35914904). These challenges result in a highly geographically heterogenous disease affecting people worldwide.

One of the most accessible and rapid approaches for the initial evaluation of mycobacterial lung disease is radiological imaging, which often provides the earliest clinically actionable information, particularly when definitive microbiologic diagnosis is delayed by low bacterial burden and prolonged culture turnaround times. Traditional radiologic interpretation relies on subjective visual assessment, which demonstrates variable interobserver agreement.^8^ Further, a formal scoring or staging system for NTM-LD and PTB does not currently exist, hindering the ability to uniformly monitor lung remodeling and disease progression. While certain imaging features have been associated with each condition, PTB characteristically presenting with upper lobe predominance and cavitation, NTM-LD with middle lobe and lingular bronchiectasis, substantial overlap exists, and prior studies have shown that these features do not offer adequate markers for reliable discrimination.^5,9^

Radiomics, the high-throughput extraction of quantitative imaging features, has emerged as a promising approach to augment clinical decision-making.^10,11^ Recent studies have applied radiomics to NTM-LD versus PTB differentiation with encouraging results. Yan and colleagues^12^ demonstrated that radiomic features extracted from cavities achieved AUC values exceeding 0.84 for differentiating NTM-LD from PTB. Similarly, consolidation-based radiomics models have achieved AUC values above 0.90.^13^ However, these lesion-specific approaches require manual delineation of individual cavities or consolidations by experienced radiologists, a labor-intensive process that limits scalability and introduces potential inter-reader variability.

Deep learning methods have also been applied to this classification task. Wang and colleagues^14^ reported an AUC of 0.71 using the BoTNet50 convolutional neural network, while 3D-ResNet models have achieved AUC values of 0.78-0.86 in a cohort from China.^15^ Although promising, these deep learning approaches function as “black boxes” with limited interpretability and require substantial computational resources, potentially hindering clinical adoption. A critical gap exists in the literature: no study has systematically evaluated whole-lung radiomic features for NTM-LD classification across geographically and demographically distinct populations. Whole-lung analysis offers various potential advantages: automated lung segmentation eliminates the need for manual lesion annotation, captures global parenchymal patterns that may not be apparent in individual lesions, and facilitates deployment in settings where expert radiologic annotation is unavailable. In this study, we evaluated whether whole-lung radiomic features could achieve robust NTM-LD classification performance across cohorts from China and the United States while offering substantially greater scalability and interpretability than existing approaches.

## STUDY DESIGN AND METHODS

### Study Population

This study utilized two geographically distinct cohorts. Dataset 1 comprised a publicly available collection of chest CT scans from patients diagnosed with pulmonary mycobacterial disease from China.^15^ This dataset represents the first comprehensive CT imaging resource combining NTM-LD and PTB cases with multispecies mycobacterial information. The dataset includes anonymized imaging data and clinical descriptors obtained prior to treatment initiation. Cases were selected with microbiologically confirmed diagnoses and CT scans of adequate quality for quantitative analysis. The final study cohort comprised 1,300 patients: 871 with culture-confirmed PTB and 429 with culture-confirmed NTM-LD, representing the largest radiomics cohort assembled for this classification task to date. Dataset 2 comprised an independent clinical NTM-LD cohort (N=173) acquired at the University of Alabama at Birmingham (UAB; IRB-300010437). (PMID: 41046132)^18^ NTM-LD patients were identified from the UAB Bronchiectasis Research Registry as having pulmonary infections with NTM but not with TB based on prior sputum cultures in the year prior. Patients with cystic fibrosis (CF) were excluded because the underlying structural lung abnormalities characteristic of CF, including diffuse bronchiectasis, mucus plugging, and progressive parenchymal destruction, can confound radiomic feature extraction and introduce disease-related imaging signatures unrelated to mycobacterial infection. Patients with extrapulmonary manifestations of NTM (i.e., skin and soft tissue) were also excluded from the study. CT scans were obtained retrospectively from the most recent visit. Clinical data was collected in October 2024. Notably, Dataset 2 did not include any TB cases, reflecting the low prevalence of active PTB in the United States; the role of this cohort in our analysis is detailed below under the experimental design.

### CT Imaging Protocol

CT imaging was performed using standardized protocols across participating centers.^15^ The scan range encompassed the entire thorax from lung apex to base, with patients positioned supine during end-inspiratory breath-hold. Scans were acquired at 512 x 512 matrix resolution on GE BrightSpeed and Canon Aquilion Prime 128 machines, with thin slices (1.0–1.25 mm) to capture fine details of the lung tissue.

### Automated Lung Segmentation

In contrast to prior radiomics studies requiring manual lesion delineation,^12,13^ we employed fully automated deep learning-based lung segmentation.^16^ This algorithm employs a U-Net architecture trained on diverse CT datasets to achieve robust segmentation across varying imaging protocols and disease presentations. Segmentation quality was visually assessed for each case.

### Radiomic Feature Extraction

Radiomic feature extraction was performed using the PyRadiomics (version 3.1.1) library.^10,11^ A total of 85 quantitative features was extracted from the segmented whole-lung parenchyma, capturing global disease burden rather than individual lesion characteristics. First-order statistics captured intensity distribution characteristics, including mean, median, standard deviation, skewness, kurtosis, and energy. Shape-based features quantified three-dimensional lung morphology, including volume, surface area, sphericity, and elongation. Texture features were derived from gray-level co-occurrence matrices (GLCM), gray-level run-length matrices (GLRLM), gray-level size zone matrices (GLSZM), and neighboring gray-tone difference matrices (NGTDM). Image preprocessing included intensity normalization and isotropic resampling to standardize voxel dimensions across different scanner protocols. Feature values were standardized using z-score normalization.

### Statistical Analysis and Model Development

Two experimental scenarios were designed to evaluate the generalizability of whole-lung radiomic features for NTM-LD classification. In Scenario 1 (Cross-Cohort External Validation), the LDA classifier was trained on Dataset 1 (China cohort; N=1,300; NTM=429, TB=871) using a 70/30 stratified train-test split (train=909, test=391). The trained model was then applied to Dataset 2 (UAB cohort; N=173, all NTM-LD) as an external validation set to evaluate whether NTM-LD radiomic signatures learned from the Chinese cohort generalize to an American NTM-LD population. In Scenario 2 (Combined Cohort Analysis), Datasets 1 and 2 were combined into a single cohort (N=1,473; NTM=602, TB=871) with a group-aware 70/30 stratified split maintaining proportional representation from each source dataset and disease class (Dataset 1 TB: train=610, test=261; Dataset 1 NTM: train=300, test=129; Dataset 2 NTM: train=121, test=52). Linear discriminant analysis (LDA) was selected as the classification algorithm owing to its computational efficiency with high-dimensional data, interpretability, and established performance in medical imaging applications.^17^ LDA offers explicit feature weights that enhance clinical interpretability.

Model performance was evaluated on the independent test set using Area under the Curve (AUC) and precision.^19,20^ Additional metrics included sensitivity, specificity, and overall accuracy. Confidence intervals for AUC were computed using bootstrap resampling with 1,000 iterations. Feature importance was assessed using SHapley Additive exPlanations (SHAP) values to identify radiomic features contributing most substantially to classification decisions.^21^ Performance was benchmarked against published results from the BoTNet50 deep learning model evaluated on the same dataset.^14^ All statistical analyses were performed using Python 3.8 with scikit-learn.^17^ Statistical significance was defined as *P* < 0.05 (two-sided).

## RESULTS

### Cohort Characteristics

Table 1 summarizes the characteristics of the two study cohorts. Dataset 1 (China cohort) comprised 1,300 patients with culture-confirmed pulmonary mycobacterial disease, including 871 PTB cases (67.0%) and 429 NTM-LD cases (33.0%), collected from multiple centers in China. This publicly available dataset contains multispecies mycobacterial CT imaging data acquired across several Chinese institutions, providing broad geographic and demographic representation within the East Asian population. Dataset 2 (UAB cohort) comprised 173 patients with culture-confirmed NTM-LD exclusively, identified from the UAB Bronchiectasis Research Registry in Birmingham, Alabama, United States. The UAB cohort was predominantly female (71.1%), non-Hispanic White (86.1%), with a mean age of 69.8 ± 11.8 years and mean BMI of 22.8 ± 5.8 kg/m². Mean percent predicted FEV₁ was 71.3% ± 23.3%, reflecting moderate airflow limitation consistent with underlying bronchiectasis. Among UAB participants, 43.4% were current or former smokers. The majority (79.8%) were symptomatic at the time of CT acquisition.

**Table 1.**
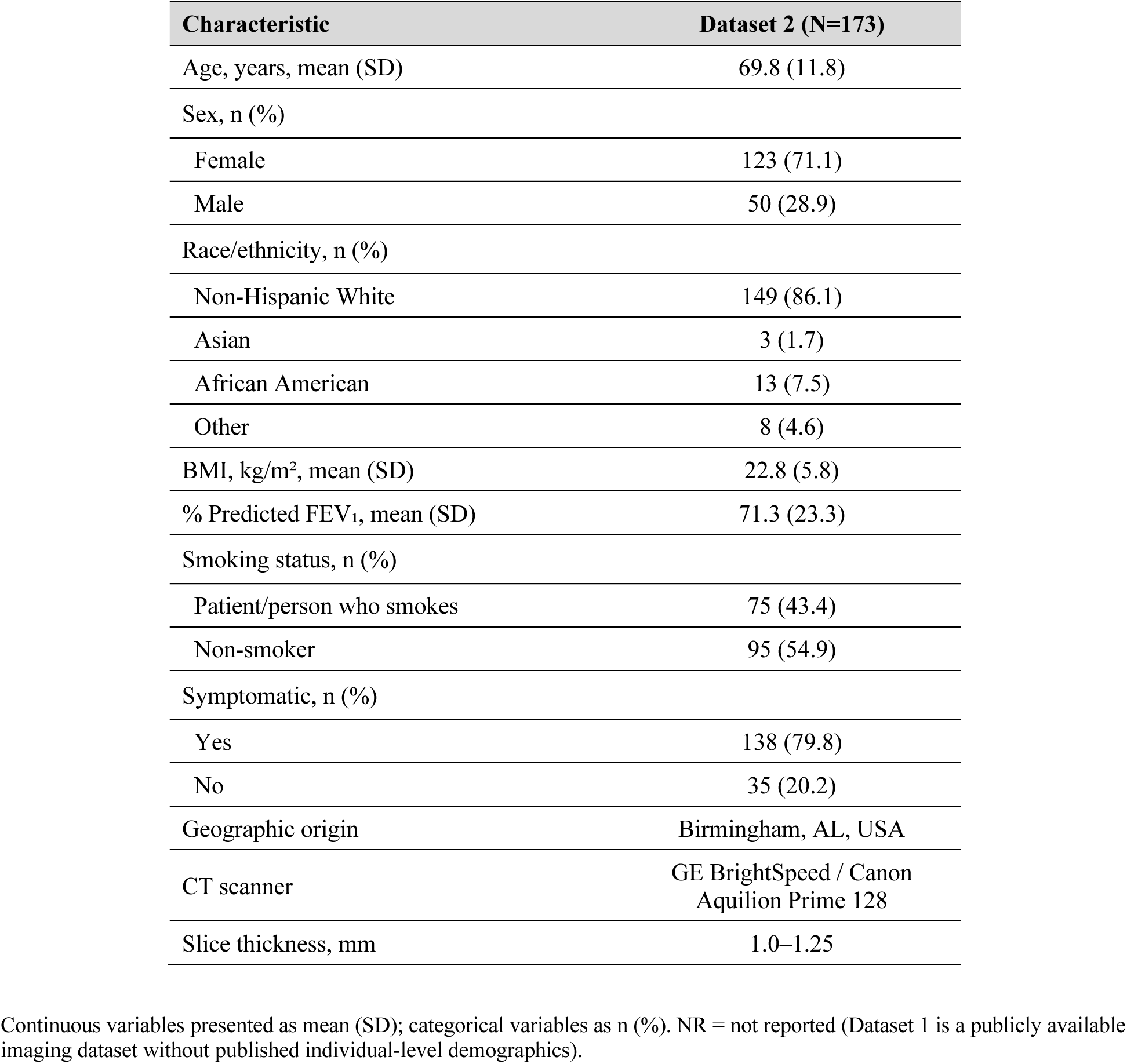
Cohort Characteristics.

### Classification Performance

The whole-lung radiomics-based LDA classifier demonstrated strong discriminative performance on the Dataset 1 internal test set (Table 2). In Scenario 1, the model achieved an AUC of 0.79 (95% CI, 0.73–0.84), sensitivity of 0.47 (95% CI, 0.39–0.56), specificity of 0.91 (95% CI, 0.88–0.94), accuracy of 0.767 (95% CI, 0.73–0.81), and an F1 score of 0.57. When applied to the external UAB cohort (Dataset 2, N=173), the model achieved an AUC of 0.94 (95% CI, 0.90–0.97), correctly detecting 162 of 173 NTM-LD cases (sensitivity 64.2%). This cross-cohort performance demonstrates that NTM-LD radiomic signatures learned from the Chinese cohort generalize effectively to an American NTM-LD population despite differences in demographics, mycobacterial species distributions, and imaging protocols.

**Table 2.** Classification Performance: Scenario 1 (Train Dataset 1, Internal Test=391) and External Validation on Dataset 2 (N=173)

| **Performance Metric** | **Scenario 1:**<br>**Internal Test**<br>**(n=391)** | **95% CI** | **External**<br>**Validation D2**<br>**(n=173)** | **95% CI** |
| --- | --- | --- | --- | --- |
| AUC | 0.79 | 0.73–0.84 | 0.91 | 0.88-0.94 |
| Sensitivity | 0.47 | 0.39–0.56 | 0.94 | 0.90–0.97 |
| Specificity | 0.91 | 0.88–0.94 | 1.00 | 1.00-1.00 |
| Accuracy | 0.77 | 0.73–0.81 | 0.94 | 0.90–0.97 |
| F1 Score | 0.57 | 0.51–0.60 | 0.97 | 0.95-0.99 |
| NTM-LD Detected | 61/173 | — | 162/173 | — |

AUC = area under the receiver operating characteristic curve; CI = confidence interval; LDA = linear discriminant analysis; External validation on Dataset 2 reports detection rate for NTM-LD; traditional specificity metrics are not applicable as Dataset 2 contains no TB cases. Dash (—) indicates metric not applicable.

In Scenario 2 (Combined Cohort; Table 3), the model achieved an AUC of 0.81 (95% CI, 0.76–0.85), sensitivity of 0.61 (95% CI, 0.54–0.69), specificity of 0.90 (95% CI, 0.87–0.94), accuracy of 0.79 (95% CI, 0.75–0.82), and an F1 score of 0.70. The improved AUC (0.81 vs. 0.79), sensitivity (0.61 vs. 0.47), and F1 score (0.70 vs. 0.57) relative to Scenario 1 indicate that incorporating NTM-LD cases from a second geographic population enriches the training data and improves the model’s ability to capture heterogeneous NTM-LD presentations.

**Table 3.** Classification Performance: Scenario 2 (Combined Cohort D1+D2, N=1,473)

| Performance Metric | Value | 95% CI |
| --- | --- | --- |
| AUC | 0.81 | 0.76–0.85 |
| Sensitivity | 0.61 | 0.54–0.69 |
| Specificity | 0.90 | 0.87–0.94 |
| Accuracy | 0.79 | 0.75–0.82 |
| F1 Score | 0.70 | 0.64–0.75 |

Combined cohort with group-aware stratified 70/30 split (D1 TB: train=610, test=261; D1 NTM: train=300, test=129; D2 NTM: train=121, test=52). Dash (—) indicates metric not applicable.

Receiver operating characteristic curve analysis demonstrated stable performance across threshold ranges, with particularly reliable behavior at high specificity levels (>0.90), where the model preserved its discriminative strength (Figure 2). Precision-recall analysis revealed consistently high precision values, remaining above 0.70 at elevated recall thresholds. Model robustness was further supported by bootstrap analysis, which produced narrow confidence intervals and demonstrated stable generalization across resampled datasets.

**Figure 1.**
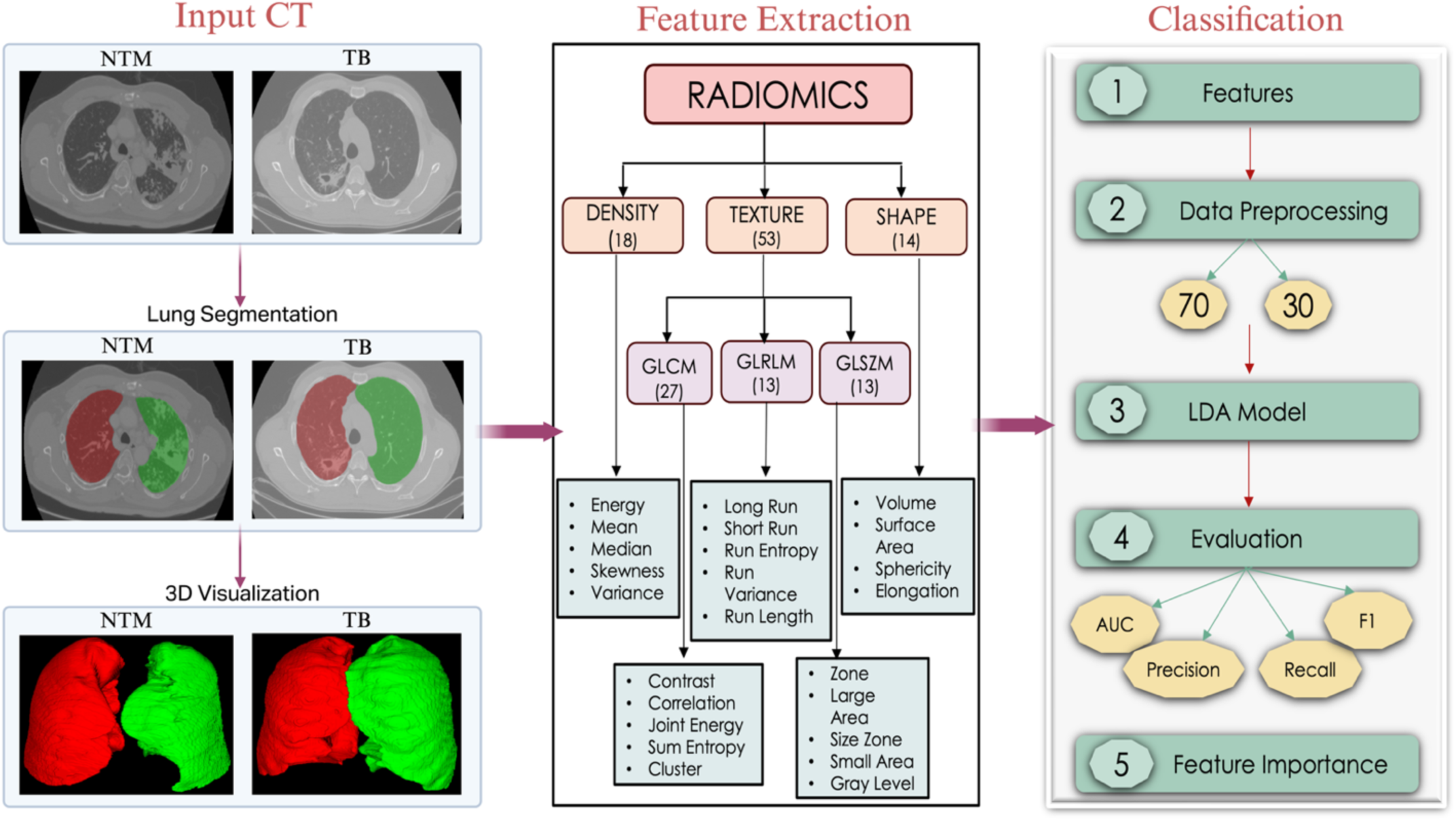
Radiomics analysis pipeline for distinguishing NTM from TB. The workflow comprises: input chest CT images from NTM and TB patient cohorts, automated lung parenchyma segmentation, quantitative radiomics feature extraction including density, texture, and shape metrics, feature selection and dimensionality reduction, Linear Discriminant Analysis (LDA) classification, and model performance evaluation with feature importance.

**Figure 2.**
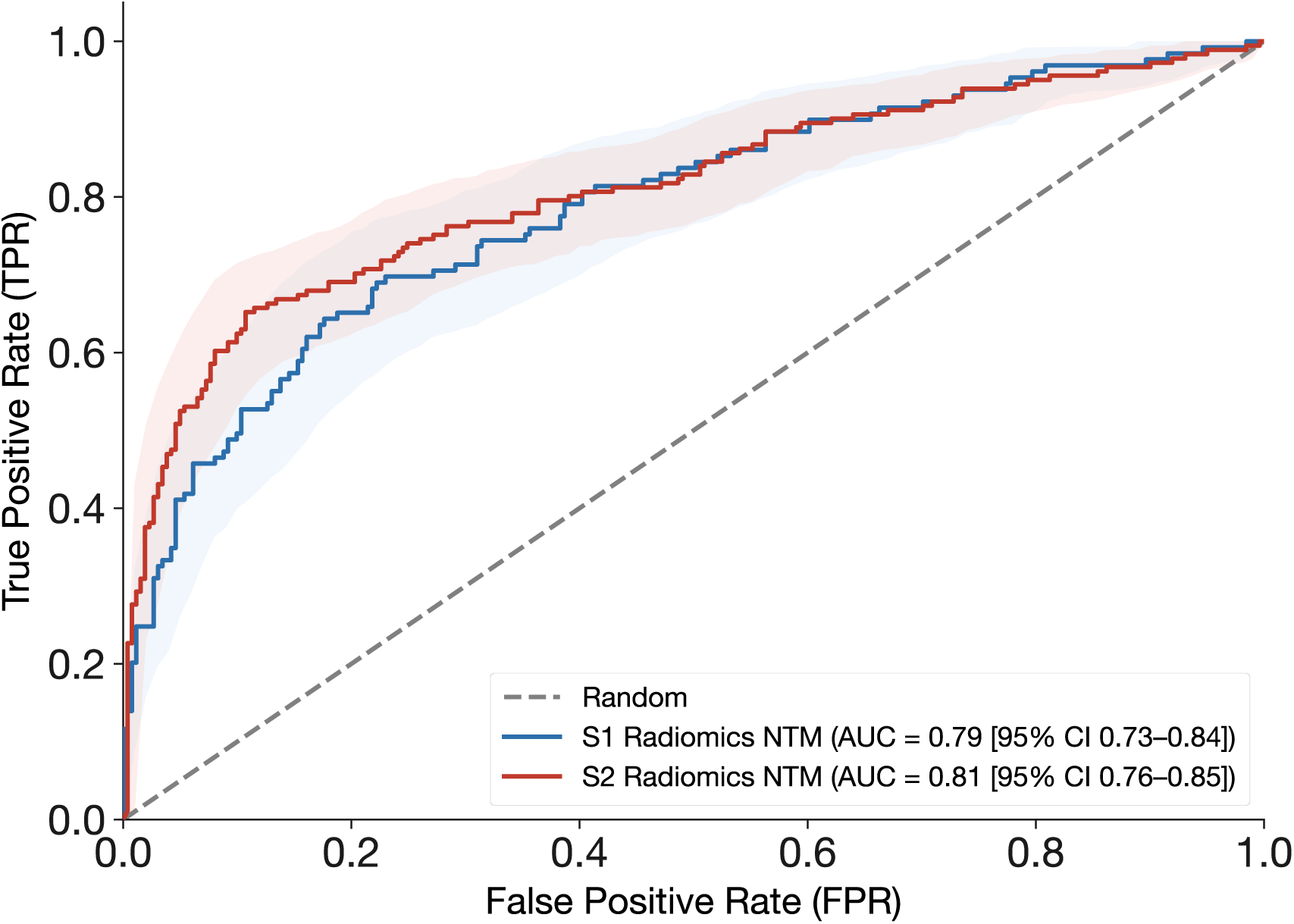
Receiver operating characteristic (ROC) curves for the whole-lung radiomics-based linear discriminant analysis classifier for Scenario 1 (S1) (Dataset 1 only) with AUC 0.79 (95% CI, 0.73–0.84) and for Scenario 2 (S2) (combined D1+D2 cohort) with AUC 0.81 (95% CI, 0.76–0.85).

### Comparison with Prior Approaches

Table 4 presents a comparison of our whole-lung approach with published methods for NTM-TB differentiation. Cavity-specific radiomics models have achieved AUC values of 0.84,^12^ and consolidation-based radiomics achieved AUC values of 0.90, but both require manual lesion-specific annotation.^13^ Our whole-lung approach achieves competitive performance (AUC = 0.79 in Scenario 1; AUC = 0.81 in Scenario 2) while requiring only automated lung segmentation, substantially reducing annotation burden and enabling application to all patients regardless of specific lesion morphology.

**Table 4.** Comparison of Classification Approaches for NTM-TB Differentiation.

| Study/<br>Approach | Method | AUC | Manual<br>Annotation | Interpretable | Cohort |
| --- | --- | --- | --- | --- | --- |
| Yan et al<sup>12</sup> | Cavity<br>radiomics + LR | 0.84 | Yes | Yes | Shandong Public Health Clinical Center<br>Jinan Infectious Disease Hospital |
| Yan et al<sup>13</sup> | Consolidation<br>radiomics | 0.90 | Yes | Yes | Shandong Public Health Clinical Center<br>Jinan Infectious Disease Hospital |
| Han et al<sup>15</sup> | BoTNet50<br>CNN | 0.71 | No | No | Mycobacterial CT Imaging Dataset<br>(Kaggle) |
| Wang et al<sup>14</sup> | 3D-ResNet | 0.78 | No | No | Tianjin Haihe Hospital, China |
| Current<br>study | Whole-lung<br>radiomics | 0.79 / 0.81 | No | Yes | D1: China (multicenter); D2: UAB<br>NTM-LD Registry (USA) |

AUC = area under the receiver operating characteristic curve; CNN = convolutional neural network; LDA = linear discriminant analysis; LR = logistic regression. Current study results shown for Scenario 1 (Dataset 1 only) and Scenario 2 (combined D1+D2 cohort).

### Feature Importance Analysis

SHAP-based feature importance analysis identified texture heterogeneity measures and morphologic descriptors as the most influential parameters for distinguishing NTM-LD from TB (Figure 3). The top-ranked features organized by radiomic class were: (1) Shape features: Voxel Volume and Mesh Volume, reflecting three-dimensional lung volume and disease-related volumetric changes; (2) GLSZM texture features: Large Area Emphasis and Zone Variance, quantifying the prevalence of large homogeneous zones and spatial heterogeneity of parenchymal texture patterns; (3) GLCM texture features: Inverse Difference Moment (Idm), Inverse Difference (Id), Autocorrelation, Difference Average, Inverse Variance, Joint Entropy, and Entropy, collectively capturing local textural uniformity, spatial intensity correlations, and gray-level complexity; (4) GLRLM texture features: Run Percentage, High Gray Level Run Emphasis, Run Length NonUniformity Normalized, Long Run Emphasis, and Run Variance, encoding the distribution of consecutive voxel runs and reflecting fine-to-coarse granularity of parenchymal patterns; and (5) First-order features: Short Run High Gray Level Emphasis, indicating small bright foci such as nodules or calcifications. Critically, 16 of these features were consistently ranked among the top 20 in both experimental scenarios, providing strong evidence for population-independent radiomic signatures of NTM-LD.

**Figure 3.**
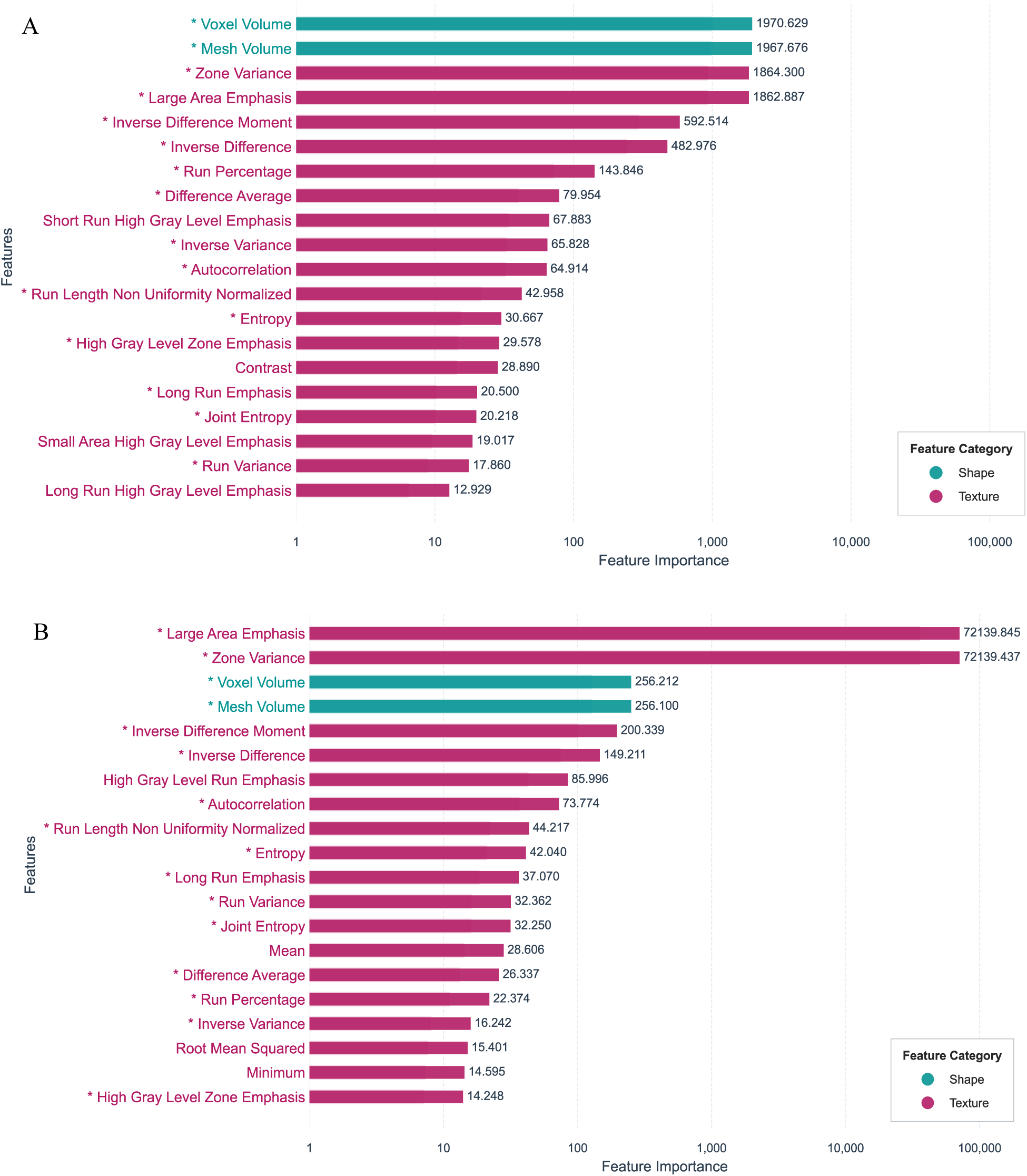
Radiomic feature contributions to differentiation between TB and NTM-LD. A, Top 20 radiomic features driving model performance in Scenario 1 (Dataset 1 only), including texture heterogeneity measures and intensity-based features; asterisks (*) denote features shared with Scenario 2 (combined D1+D2 cohort). B, Top 20 radiomic features in Scenario 2 (combined D1+D2 cohort); asterisks (*) denote features shared with Scenario 1 (Dataset 1 only).

### Cross-Cohort Validation with Shared Radiomic Features

We evaluated the generalizability of discriminative radiomic features across geographically distinct cohorts. We performed cross-cohort validation using a shared feature set derived from both datasets. Each dataset was partitioned into training (70%) and testing (30%) subsets with stratified sampling. Feature selection was performed independently within each training partition, and the intersection yielded 16 shared radiomic features common to both geospatial cohorts (Table 5). These 16 shared features spanned four radiomic classes. Shape descriptors (VoxelVolume, MeshVolume) captured three-dimensional lung morphology. GLCM texture features (Idm, Id, Autocorrelation, JointEntropy, DifferenceAverage, InverseVariance, Entropy) encoded local gray-level spatial dependencies and textural complexity. GLRLM features (HighGrayLevelRunEmphasis, RunLengthNonUniformityNormalized, LongRunEmphasis, RunVariance, RunPercentage) characterized the distribution and heterogeneity of consecutive voxel intensity runs. GLSZM features (LargeAreaEmphasis, ZoneVariance) quantified the size and variability of connected homogeneous zones. The predominance of texture-based features across both scenarios suggests that NTM-LD and TB produce fundamentally different global patterns of parenchymal spatial heterogeneity detectable through whole-lung analysis.

**Table 5.** Sixteen Shared Radiomic Features Identified Across Both Experimental Scenarios.

| Radiomic Class | Feature | Description | Importance Rank (16-Feature Model) |
| --- | --- | --- | --- |
| Shape | VoxelVolume | Total volume of segmented lung voxels | 3 |
| Shape | MeshVolume | Volume computed from triangular mesh of lung surface | 4 |
| GLCM (Texture) | Autocorrelation | Spatial correlation of gray-level co-occurrences | 11 |
| GLCM (Texture) | Idm (Inverse Difference Moment) | Local homogeneity of gray-level distribution | 13 |
| GLCM (Texture) | Id (Inverse Difference) | Smoothness of gray-level transitions between voxels | 8 |
| GLCM (Texture) | JointEntropy | Randomness in joint gray-level distribution of voxel pairs | 14 |
| GLCM (Texture) | DifferenceAverage | Mean absolute difference between paired gray levels | 16 |
| GLCM (Texture) | InverseVariance | Inverse-weighted variance of gray-level differences | 10 |
| GLCM (Texture) | Entropy | Uncertainty/complexity in gray-level spatial distribution | 15 |
| GLRLM (Texture) | HighGrayLevelRunEmphasis | Concentration of high-intensity consecutive voxel runs | 12 |
| GLRLM (Texture) | RunLengthNonUniformityNormalized | Heterogeneity of run lengths across the image | 6 |
| GLRLM (Texture) | LongRunEmphasis | Predominance of long consecutive voxel runs | 7 |
| GLRLM (Texture) | RunVariance | Dispersion of run length values | 9 |
| GLRLM (Texture) | RunPercentage | Fraction of voxels that form runs relative to total | 5 |
| GLSZM (Texture) | LargeAreaEmphasis | Predominance of large, connected zones of similar intensity | 1 |
| GLSZM (Texture) | ZoneVariance | Dispersion of zone sizes across the image | 2 |

GLCM = gray-level co-occurrence matrix; GLRLM = gray-level run-length matrix; GLSZM = gray-level size-zone matrix. Importance rank derived from the 16-feature reduced LDA model (Scenario 1). Features are grouped by radiomic class and ordered by importance within each class.

An LDA classifier trained and evaluated on Dataset 1 using only these 16 features retained 94.4% of the full model’s discriminative performance (AUC 0.74 [95% CI, 0.69–0.80] vs. 0.79 [95% CI, 0.73–0.84]). Notably, the reduced model surpassed the full model in specificity (0.95 [95% CI, 0.93–0.98] vs. 0.91 [95% CI, 0.88–0.94]) and positive predictive value (0.82 [95% CI, 0.73–0.91] vs. 0.73 [95% CI, 0.63–0.82]), with accuracy of 0.78 (95% CI, 0.74–0.82). The sensitivity trade-off (0.43 [95% CI, 0.35–0.52] vs. 0.65 [95% CI, 0.58–0.72]) yielded an F1 of 0.57.

The 16 texture and shape features were predominantly texture features, GLCM and GLRLM descriptors encoding spatial gray-level dependencies and run-length distributions, retaining 94.4% of discriminative capacity with overlapping confidence intervals (AUC 0.74 [95% CI, 0.69–0.80] vs. 0.79 [95% CI, 0.73–0.84]). The concurrent improvement in specificity suggests that feature reduction functions as implicit regularization, isolating high-confidence TB-specific structural patterns while attenuating overfitting to cohort-specific variance. The reduction in sensitivity likely reflects exclusion of features that captured heterogeneous NTM presentations in individual cohorts but did not replicate across populations, consistent with the greater radiographic variability characteristic of NTM disease.

## DISCUSSION

In this study, we demonstrated that whole-lung radiomic features extracted from chest CT scans enable robust classification of NTM-LD across geographically and demographically distinct populations using an interpretable machine learning approach that requires no manual lesion annotation. The radiomics-based LDA classifier achieved strong discriminative performance both in a single-cohort setting (AUC = 0.79) and when applied cross-nationally (external validation AUC = 0.94), while a combined cohort model further improved classification (AUC = 0.81). These results exceeded published deep learning methods^14^ while approaching performance of lesion-specific radiomics models^12,13^ all without requiring the labor-intensive manual delineation of individual cavities or consolidations.

A key finding is the strong generalizability of NTM-LD radiomic signatures across populations. When the model trained exclusively on the Chinese cohort (Dataset 1) was applied to the American UAB cohort (Dataset 2), it achieved an AUC of 0.94, correctly identifying 162 of 173 NTM-LD cases. This cross-cohort performance is notable because the two populations differ substantially in demographics, geographic origin, likely mycobacterial species distributions, and imaging protocols. The fact that whole-lung radiomic features captured NTM-LD patterns that transcend these sources of variability suggests that NTM-LD produces fundamental, population-independent alterations in global lung parenchymal texture and morphology. The identification of 16 radiomic features consistently ranked among the top 20 in both experimental scenarios further reinforces this conclusion. The discriminative features identified through SHAP capture global parenchymal patterns consistent with known pathophysiologic differences between TB and NTM.^5,6^ TB characteristically produces more extensive tissue destruction with coarse, heterogeneous parenchymal changes, whereas NTM lung disease, particularly that caused by Mycobacterium avium complex, may demonstrate more diffuse or nodular involvement with relatively preserved parenchymal architecture.^1,23^ The predominance of texture-driven and intensity-driven features in our classification model quantitatively captures these radiologic distinctions at a whole-lung level. The importance of shape-based metrics further suggests that the overall geometric configuration of affected lung contributes meaningfully to differentiation between these disease entities.

Deep learning methods, while achieving moderate performance (AUC = 0.71-0.78),^14,15^ function as “black boxes” that provide limited insight into classification rationale. Our feature-based radiomics model with explicit SHAP-based feature importance quantification identifies specific imaging characteristics driving predictions. The predominance of texture heterogeneity features (zone variance, inverse difference moment) aligns with known differences in parenchymal destruction patterns between TB and NTM,^5,23^ providing clinical face validity that may enhance physician trust and adoption. Importantly, cross-cohort validation demonstrated that a reduced set of 16 shared features retained 94.4% of the full model’s performance while improving specificity and positive predictive value. These shared features were predominantly texture-based (GLCM and GLRLM), capturing spatial gray-level heterogeneity across the lung parenchyma. This supports that global texture patterns are robust and reproducible markers of TB versus NTM across populations. The improved specificity with fewer features suggests implicit regularization, reducing cohort-specific noise, whereas the decrease in sensitivity likely reflects exclusion of features associated with the more heterogeneous presentation of NTM disease.

Recent work by Biciusca and colleagues^24^ highlighted the challenge of differentiating radiologic features among slow-growing NTM species, finding that *M. intracellulare* exhibited a tendency toward higher overall radiologic severity scores compared with other species. Such species-level heterogeneity within NTM infections underscores the complexity of this diagnostic challenge. Likewise, heterogeneity in the etiology of NTM-LD may lead to distinct patterns of disease progression that are currently unknown due to the lack a formal scoring/staging system and standardized analytic tools. Our model’s ability to distinguish NTM as a group from TB, despite this intra-group variability, suggests that quantitative radiomic features capture fundamental differences in parenchymal involvement patterns between these mycobacterial categories. The high specificity (0.91) of our classifier warrants particular attention, as it indicates robust ability to correctly classify non-NTM cases and supports confident clinical decision-making.^4^ A highly specific classifier minimizes misclassification, ensuring that patients with NTM-LD are correctly identified and can receive timely, appropriate therapeutic regimens.^2^ The precision-recall characteristics of our model support its utility in clinical workflows where confident NTM-LD identification is needed alongside microbiologic confirmation.

Our study has several limitations. First, our analysis was retrospective, utilizing a publicly available dataset and a clinical registry cohort, both with inherent selection biases given the cohort’s location. Prospective and longitudinal validation in diverse clinical settings is necessary before clinical implementation. Second, Dataset 2 did not include TB cases, precluding traditional sensitivity/specificity evaluation on this cohort for NTM vs. TB discrimination; its role was to validate the generalizability of NTM-LD radiomic signatures rather than to serve as a full NTM vs. TB discrimination test. Third, we analyzed whole-lung radiomic features without lesion-specific analysis; a combined approach incorporating both whole-lung and lesion-level features might further improve classification accuracy. Fourth, while our model outperformed deep learning approaches evaluated on the same dataset, more advanced neural network architectures may achieve superior performance. Finally, clinical integration would require validation of workflow feasibility, comparison with expert radiologist interpretations, and assessment of the model’s impact on clinical decision-making.

## CONCLUSION

Whole-lung CT radiomics enables interpretable machine learning-based classification of NTM-LD across geographically and demographically distinct populations without requiring manual lesion annotation. This automated approach achieves performance exceeding deep learning methods and approaching that of lesion-specific radiomics models while offering substantially greater scalability and interpretability. The identification of 16 shared radiomic features that generalize across Chinese and American cohorts suggests that NTM-LD produces distinct, reproducible, population-independent parenchymal signatures detectable through quantitative imaging analysis. This lightweight, interpretable approach may facilitate NTM-LD detection when microbiologic confirmation is pending, particularly in diverse clinical settings. Future directions include prospective multicenter validation, integration with clinical and laboratory data including biomarkers, comparison with expert radiologist interpretations, and combination with lesion-specific features to potentially enhance discriminative performance. ^25^

## Data Availability

Data are available through two sources: (1) publicly in Kaggle at https://www.kaggle.com/datasets/damianhan/dicom-dataset/data, and (2) through the University of Alabama at Birmingham institutional repository, which requires IRB approval.

## Other contributions

The authors thank Han and colleagues for making the mycobacterial CT imaging dataset publicly available, enabling this research.

